# Predictors of parents’ intention to vaccinate their children against the COVID-19 in Greece: a cross-sectional study

**DOI:** 10.1101/2021.09.27.21264183

**Authors:** Petros Galanis, Irene Vraka, Olga Siskou, Olympia Konstantakopoulou, Aglaia Katsiroumpa, Ioannis Moisoglou, Daphne Kaitelidou

**Affiliations:** Clinical Epidemiology Laboratory, Faculty of Nursing, National and Kapodistrian University of Athens, Athens, Greece; Department of Radiology, P & A Kyriakou Children’s Hospital, Athens, Greece; Center for Health Services Management and Evaluation, Faculty of Nursing, National and Kapodistrian University of Athens, Athens, Greece; Pulmonary Clinic, General Hospital of Lamia, Lamia, Greece

**Keywords:** COVID-19, parents, children, intention, willingness

## Abstract

**Background:** Parents’ intention to vaccinate their children against the COVID-19 is envisaged as critical strategy to control the pandemic.

**Objective:** To investigate the intention of parents to vaccinate their children against the COVID-19 and the factors influencing this intention.

**Methods:** We conducted an online cross-sectional study in Greece and we collected data during the first week of September 2021. A convenience sample was used by collecting questionnaires through social media. Our study population included adult parents with children aged 12-17 years who were eligible for a COVID-19 vaccine.

**Results:** Study population included 813 parents with a mean age of 42.3 years. Among parents, 36% reported that they will vaccinate their children against the COVID-19, 33.5% denied vaccination and 30.5% were undecided. Concerns about the safety, effectiveness and side effects of COVID-19 vaccines were the most important reasons for decline of COVID-19 vaccination. Parents who took the flu vaccine in 2020 and those who had more knowledge and fewer concerns about COVID-19 vaccines had a greater probability to vaccinate their children against the COVID-19. Increased self-perceived severity of COVID-19, and increased trust in COVID-19 vaccines and the government regarding the information about the COVID-19 vaccines were associated with parents’ intention to vaccinate their children. However, increased knowledge regarding COVID-19 was associated with decreased intention of parents to vaccinate their children.

**Conclusions:** Parents’ intention to vaccinate their children against the COVID-19 was low. Our findings could contribute to the development of target strategies to implement adherence to COVID-19 vaccination campaigns.

## Introduction

Worldwide, mass vaccination against the Coronavirus disease 2019 (COVID-19) is envisaged as critical strategy to control the pandemic (Graham, 2020). Severe illness and mortality from the COVID-19 are quite low for children but a sharp rise in infections is driven by the delta variant (Tanne, 2021). Moreover, the percentage of the population that needs to be vaccinated against the COVID-19 to achieve herd immunity is now approaching 80% (Bartsch et al., 2020). Thus, the decision of parents to vaccinate their children is crucial to limit the negative consequences of the pandemic.

In May 2021, European, USA and Canadian regulators were the first to approve a COVID-19 vaccine for 12 to 17-year-olds (European Medicines Agency, 2021). The rollout started at different times in different countries. For instance, the rollout started immediately in the USA and by the end of July, 42% of children aged 12-17 years had already received the first dose of a COVID-19 vaccine (BBC, 2021). Since the beginning of June, Canada, China, Israel and several European countries such as France, Germany, and Italy have followed the example of the USA.

According to a meta-analysis (Galanis et al., 2021), parents’ intention to vaccinate their children against the COVID-19 ranges from 29% to 72.7%, while the overall proportion of parents that intend to vaccinate their children is 56.8%. Parents’ willingness to vaccinate their children against COVID-19 is affected by several factors such as gender (Goldman et al., 2020; Kelly et al., 2021; Montalti et al., 2021; Yigit et al., 2021), age of parents and children (Goldman et al., 2020; Kelly et al., 2021; Montalti et al., 2021; Skjefte et al., 2021; Szilagyi et al., 2021), socio-economic status (Brandstetter et al., 2021; Hetherington et al., 2021; Kelly et al., 2021; Montalti et al., 2021; Skjefte et al., 2021; Szilagyi et al., 2021; Wang et al., 2021; Yigit et al., 2021), race (Bell et al., 2020; Kelly et al., 2021; Scherer et al., 2021; Teasdale et al., 2021), attitudes toward vaccination (Goldman et al., 2020; Kelly et al., 2021; Ruggiero et al., 2021; Skjefte et al., 2021; Yilmaz and Sahin, 2021; Zhang et al., 2020), and information about the COVID-19 (Brandstetter et al., 2021; Montalti et al., 2021; Zhang et al., 2020).

As mentioned above, several studies have investigated parents’ intention to vaccinate their children against the COVID-19, but only four studies were conducted during 2021 (Scherer et al., 2021; Szilagyi et al., 2021; Teasdale et al., 2021; Yilmaz and Sahin, 2021) and none of these studies was conducted in Europe. In addition, all studies investigated parental intention before a COVID-19 vaccine received official approval to be administered to children by the authorities. Therefore, the studies to date have assessed parental attitudes towards a hypothetical safe and effective COVID-19 vaccine. Moreover, these studies captured parents’ opinions during a period that it was still unknown whether and when COVID-19 vaccines for children would be available.

Therefore, our study investigated the intention of parents to vaccinate their children against the COVID-19 with a proven safe and effective vaccine that has received official approval and can now be administered to children after parental consent. In addition, we also investigated the factors that influence parents’ intention to vaccinate their children.

## Methods

### Study design and participants

We conducted an online cross-sectional study in Greece and we collected data during the first week of September 2021. The Greek government is offering a COVID-19 vaccine free of charge to all children aged 15-17 years from 15 July 2021 and from 30 July to all children aged 12-14 years. Thus, our study population included adult parents with children aged 12-17 years who were eligible for a COVID-19 vaccine but had not yet received it. A convenience sample was used by collecting questionnaires through social media. Specifically, we used google forms to create the online version of the study questionnaire which we then posted on social media. Parents were informed about the purpose and methodology of the study and consented to participate.

### Independent variables

Independent variables in our study included socio-demographic data of parents and attitudes towards vaccination and COVID-19 pandemic. In particular, socio-demographic data included the following: gender, parents’ age, marital status (singles, married, divorced, and widowed), educational level (elementary school, high school, University degree, MSc/PhD degree), working on healthcare facilities, self-perceived financial status and health status, parents’ chronic disease, parents’ previous COVID-19 diagnosis, family/friends with previous COVID-19 diagnosis, and living with elderly people or vulnerable groups during the COVID-19 pandemic. Financial status and health status were measured with a five-point Likert scale from 0 (“very poor”), to 4 (“very good”).

Additionally, we measured children’s complete vaccination history, parents’ COVID-19 vaccination uptake and parents’ seasonal influenza vaccination in 2020 through “yes/no” answers. Reasons for decline of parents to vaccinate their children against the COVID-19 were also recorded.

We recorded parents’ attitudes towards vaccination and COVID-19 pandemic through the following variables: self-perceived knowledge regarding COVID-19 and COVID-19 vaccines, self-perceived severity of COVID-19, trust in COVID-19 vaccines, concerns about the side effects of COVID-19 vaccination, the role of vaccination in the promotion of public health, and trust in the government, scientists, and family doctors regarding the information about the COVID-19 vaccines. All these variables were recorded on a scale from 0 to 10 with higher values indicate higher level of trust, concerns, knowledge, etc.

### Outcome

Outcome variable was parents’ intention to vaccinate their children against the COVID-19 measured with “yes/no/I don’t know” answers. For statistical reasons, we considered parents’ intention as a dichotomous variable merging the “no/I don’t know” categories.

### Ethical issues

The Ethics Committee of Department of Nursing, National and Kapodistrian University of Athens approved the study protocol (reference number; 370, 02-09-2021).

### Statistical analysis

Categorical variables are presented as numbers (percentages) and continuous variables as mean (standard deviation). Regarding self-perceived financial status and health status due to low number of parents in some categories, we merged the following categories: “very poor” and “poor”; “good” and “very good”. We used univariate and multivariate logistic regression models to investigate the relationship between the independent variables and parents’ intention to vaccinate their children against the COVID-19. First, we performed univariate analysis and independent variables with a p-value <0.2 were included in a multivariate logistic regression model with backward stepwise method to eliminate confounding. In multivariate logistic regression analysis, p-values < 0.05 were considered significant. Statistical analysis was performed with the Statistical Package for Social Sciences software (IBM Corp. Released 2012. IBM SPSS Statistics for Windows, Version 21.0. Armonk, NY: IBM Corp.).

## Results

Detailed socio-demographic profile of parents is shown in Table 1. Study population included 813 parents with a mean age of 42.3 years. The majority of parents was mothers (76.1%) and married (88.1%). Among parents, 10.2% were diagnosed with the COVID-19 and 56.3% had family/friends with a previous COVID-19 diagnosis. Most parents reported a moderate/good financial status (83.8%) and a good/very good health status (79.9%).

**Table 1.** Socio-demographic profile of parents.

| **Characteristics** | **N** | **%** |
| --- | --- | --- |
| Gender |  |  |
| Mothers | 619 | 76.1 |
| Fathers | 194 | 23.9 |
| Parents' age (years)<sup>a</sup> | 42.3 | 7.4 |
| Marital status |  |  |
| Singles | 32 | 3.9 |
| Married | 716 | 88.1 |
| Divorced | 61 | 7.5 |
| Widowed | 4 | 0.5 |
| Educational level |  |  |
| Elementary school | 10 | 1.2 |
| High school | 162 | 19.9 |
| University degree | 641 | 78.8 |
| MSc/PhD degree |  |  |
| No | 468 | 57.6 |
| Yes | 345 | 42.4 |
| Working on healthcare facilities |  |  |
| No | 431 | 53.0 |
| Yes | 382 | 47.0 |
| Self-perceived financial status |  |  |
| Very poor | 15 | 1.8 |
| Poor | 76 | 9.3 |
| Moderate | 459 | 56.5 |
| Good | 222 | 27.3 |
| Very good | 41 | 5.0 |
| Self-perceived health status |  |  |
| Very poor | 3 | 0.4 |
| Poor | 25 | 3.1 |
| Moderate | 135 | 16.6 |
| Good | 410 | 50.4 |
| Very good | 240 | 29.5 |
| Parents' chronic disease |  |  |
| No | 653 | 80.3 |
| Yes | 160 | 19.7 |
| Previous COVID-19 diagnosis |  |  |
| No | 730 | 89.8 |
| Yes | 83 | 10.2 |
| Family/friends with previous COVID-19 diagnosis |  |  |
| No | 355 | 43.7 |
| Yes | 458 | 56.3 |
| Living with elderly people or vulnerable groups during the COVID-19 pandemic |  |  |
| No | 592 | 72.8 |
| Yes | 221 | 27.2 |

<sup>a</sup> mean, standard deviation

We presented parents’ attitudes towards vaccination and COVID-19 pandemic in Table 2. Most parents had been vaccinated against the COVID-19 (84.1%), while about half had been vaccinated against influenza (55%). Among parents, 36% reported that they will vaccinate their children against the COVID-19, 33.5% denied vaccination and 30.5% were undecided. Concerns about the safety, effectiveness and side effects of COVID-19 vaccines were the most important reasons for decline of COVID-19 vaccination. Parents reported a high level of knowledge regarding COVID-19 and COVID-19 vaccines but a lower level of trust in COVID-19 vaccines. Trust in family doctors regarding the information about the COVID-19 vaccines was high but it was lower in scientists and even lower in the government.

**Table 2.**
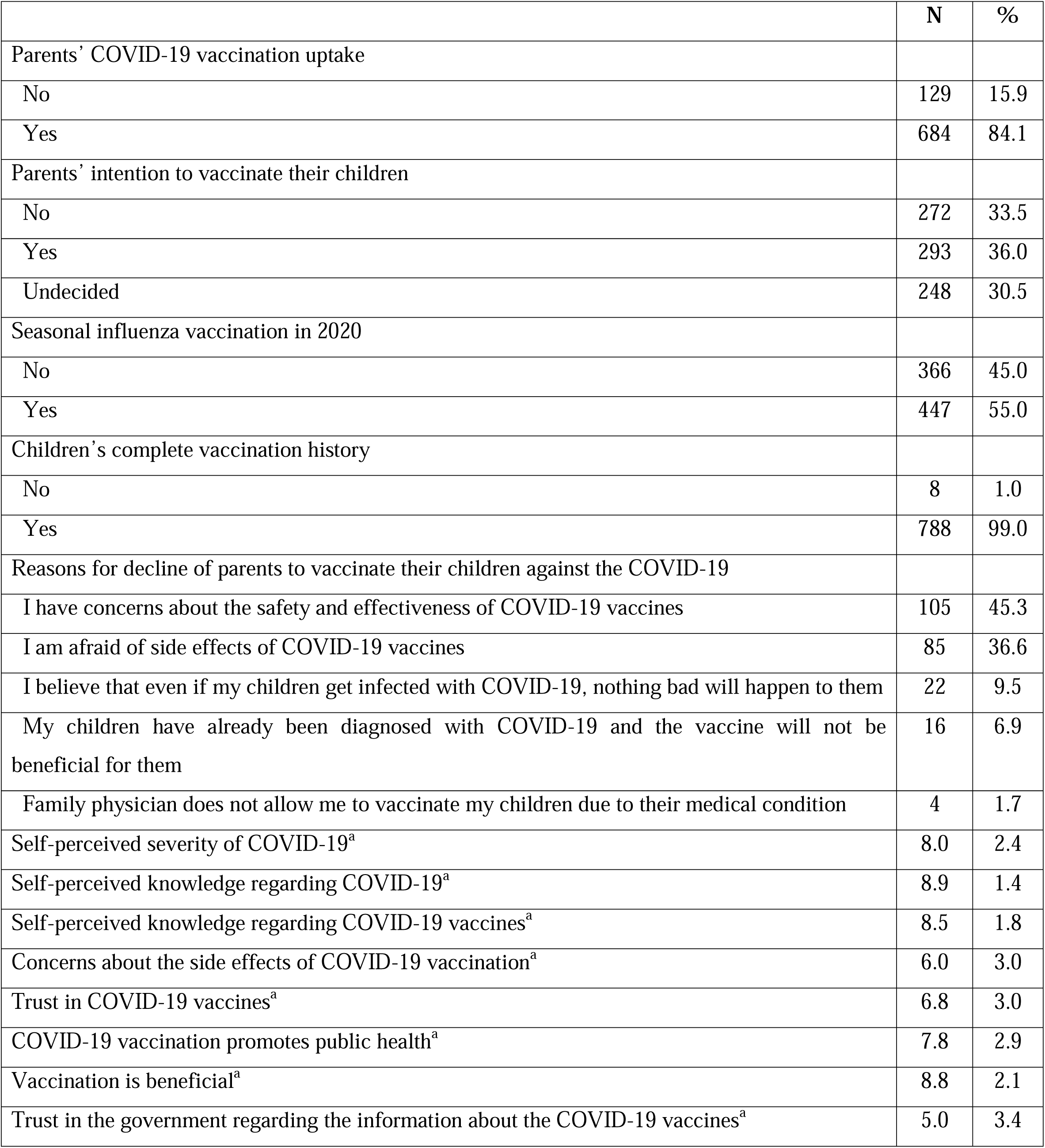
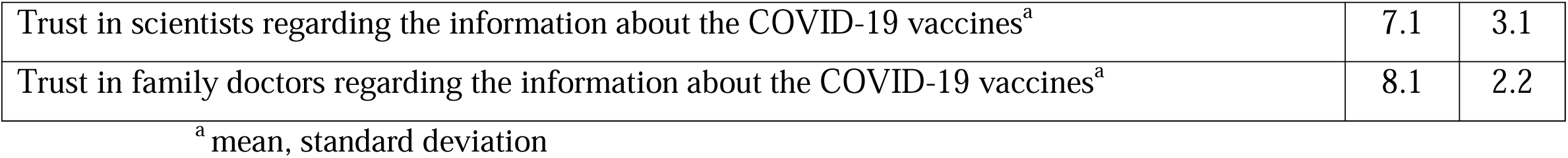
Parents’ attitudes towards vaccination and COVID-19 pandemic.

Detailed results for univariate and multivariate logistic regression analysis are presented in Table 3. Socio-demographic characteristics that were associated with parents’ intention to vaccinate their children against the COVID-19 were age, marital status, and parents’ seasonal influenza vaccination history. In particular, increased parents’ age was related with increased parents’ intention to accept a COVID-19 vaccine for their children. Additionally, singles/widowed/divorced parents and those who took the flu vaccine in 2020 were more likely to vaccinate their children against the COVID-19. Parental behavior towards vaccination and COVID-19 pandemic played an important role in parents’ decision to vaccinate their children. In more detail, parents who had more knowledge and fewer concerns about COVID-19 vaccines had a greater probability to vaccinate their children against the COVID-19. However, increased knowledge regarding COVID-19 was associated with decreased intention of parents to vaccinate their children. On the other hand, increased self-perceived severity of COVID-19, and increased trust in COVID-19 vaccines and the government regarding the information about the COVID-19 vaccines was associated with parents’ intention to vaccinate their children.

**Table 3.** Univariate and multivariate logistic regression analysis with parents’ intention to vaccinate their children as the dependent variable (reference: parents who refuse a COVID-19 vaccine or are undecided).

| Variable | Unadjusted OR (95% CI) | P-value | Adjusted OR (95% CI)<sup>a</sup> | P-value |
| --- | --- | --- | --- | --- |
| Gender (fathers vs. mothers) | 1.34 (0.96 – 1.86) | 0.08 | NS |  |
| Parents' age (years) | 1.06 (1.04 – 1.09) | <0.001 | 1.09 (1.07 – 1.13) | <0.001 |
| Marital status (singles/widowed/divorced vs. married) | 1.35 (0.88 – 2.08) | 0.17 | 1.96 (1.06 – 3.61) | 0.03 |
| Educational level (University degree vs. high school) | 1.60 (1.09 – 2.33) | 0.02 | NS |  |
| MSc/PhD degree (yes vs. no) | 0.99 (0.74 – 1.33) | 0.96 | NS |  |
| Working on healthcare facilities (yes vs. no) | 1.10 (0.82 – 1.46) | 0.53 | NS |  |
| Self-perceived financial status<sup>b</sup> |  |  | NS |  |
| Good/very good | 1.80 (1.07 – 3.01) | 0.03 |  |  |
| Moderate | 1.30 (0.79 – 2.13) | 0.29 |  |  |
| Very poor/poor | 1 (reference) |  |  |  |
| Self-perceived health status<sup>b</sup> |  |  | NS |  |
| Good/very good | 0.85 (0.39 – 1.85) | 0.67 |  |  |
| Moderate | 0.94 (0.41 – 2.16) | 0.88 |  |  |
| Very poor/poor | 1 (reference) |  |  |  |
| Parents' chronic disease (yes vs. no) | 1.08 (0.76 – 1.55) | 0.69 | NS |  |
| Previous parents' COVID-19 diagnosis (no vs. yes) | 1.64 (0.98 – 2.73) | 0.06 | NS |  |
| Family/friends with COVID-19 disease (no vs. yes) | 1.30 (0.97 – 1.73) | 0.08 | NS |  |
| Living with elderly people or vulnerable groups during the COVID-19 pandemic (yes vs. no) | 1.12 (0.82 – 1.55) | 0.48 | NS |  |
| Parents' COVID-19 vaccination uptake (yes vs. no) | 14.82 (6.44 – 34.10) | <0.001 | NS |  |
| Parents' seasonal influenza vaccination in 2020 (yes vs. no) | 2.74 (2.02 – 3.72) | <0.001 | 1.49 (1.02 – 2.20) | 0.04 |
| Self-perceived severity of COVID-19 | 1.52 (1.38 – 1.67) | <0.001 | 1.21 (1.05 – 1.39) | 0.007 |
| Self-perceived knowledge regarding COVID-19 | 1.22 (1.08 – 1.37) | 0.001 | 0.61 (0.47 – 0.78) | <0.001 |
| Self-perceived knowledge regarding COVID-19 vaccines | 1.53 (1.36 – 1.72) | <0.001 | 1.44 (1.15 – 1.80) | 0.001 |
| Concerns about the side effects of COVID-19 vaccination | 0.75 (0.71 – 0.80) | <0.001 | 0.91 (0.85 – 0.98) | 0.01 |
| Trust in COVID-19 vaccines | 1.82 (1.64 – 2.02) | <0.001 | 1.39 (1.18 – 1.65) | <0.001 |
| COVID-19 vaccination promotes public health | 1.91 (1.67 – 2.18) | <0.001 | NS |  |
| Vaccination is beneficial | 1.83 (1.56 – 2.14) | <0.001 | NS | 0.01 |
| Trust in the government regarding the information about the COVID-19 vaccines | 1.31 (1.25 – 1.38) | <0.001 | 1.07 (1.00 – 1.15) | 0.049 |
| Trust in scientists regarding the information about the COVID-19 vaccines | 1.42 (1.32 – 1.52) | <0.001 | NS |  |
| Trust in family doctors regarding the information about the COVID-19 vaccines | 1.52 (1.37 – 1.68) | <0.001 | NS |  |

An odds ratio <1 indicates a negative association, while an odds ratio >1 indicates a positive association.
CI: confidence interval; NS: not selected by the backward elimination procedure in the multivariate logistic regression analysis with a significance level set at 0.05; OR: odds ratio
<sup>a</sup> R<sup>2</sup> for the final multivariate model was 48.1%
<sup>b</sup> Due to low number of parents, we merged the following categories: “very poor” and “poor”; “good” and “very good”

## Discussion

Our study is the first that investigate parents’ willingness to vaccinate their children against the COVID-19 after a COVID-19 vaccine has received official approval from the authorities. Almost one in three parents (36%) stated that they would vaccinate their children. Only one study in Turkey (Yigit et al., 2021) reported a percentage (29%) lower than the one we found. Several other studies worldwide found that parents’ intention to vaccinate their children is higher than our study and ranges from 36.7% to 72.7% (Galanis et al., 2021). In particular, studies in Europe (United Kingdom, Germany and Italy) found that the proportion of parents’ that intend to vaccinate their children ranges from 48% to 60.4% (Bell et al., 2020; Brandstetter et al., 2021; Montalti et al., 2021). The higher proportion in the previous studies may be due to the fact that parents responded in the hypothetical scenario that there would be a safe and effective COVID-19 vaccine for children, whereas in our study parents knew that an approved vaccine is now available. However, it is encouraging that the majority of parents in our study (84.1%) had been vaccinated against the COVID-19 which shows a positive attitude towards COVID-19 vaccination.

Regarding the factors that influence parents’ intention to vaccinate their children, we found that parents who had a higher level of knowledge and fewer concerns about COVID-19 vaccines were more likely to vaccinate their children. This finding is confirmed by the literature since confidence in COVID-19 vaccine safety and effectiveness is one of the strongest predictors of vaccine acceptance (Ruggiero et al., 2021; Skjefte et al., 2021; Yilmaz and Sahin, 2021; Zhang et al., 2020). Moreover, confidence in COVID-19 vaccine safety and effectiveness shows a great variation among countries affecting vaccine acceptance (Skjefte et al., 2021). In particular, a high likelihood of COVID-19 vaccine acceptance in middle-income countries (e.g. South Africa, India and Brazil) was found (Lazarus et al., 2020). Positive COVID-19 vaccine attitude of people in middle-income countries may be due to the fact that various infectious diseases threaten individuals’ lives and public health in these countries. By providing valid and sufficient information on COVID-19 vaccine safety, parents will understand the importance of vaccination and the uptake rate for the children will be improved (Teasdale et al., 2021)

Our multivariate analysis revealed a positive relationship between parents’ self-perceived severity of COVID-19 and their intention to vaccinate their children. Several studies confirm our finding that risk perception of COVID-19 is a strong determinant of parents’ COVID-19 vaccine acceptance (Kelly et al., 2021; Skjefte et al., 2021; Xu et al., 2021; Yigit et al., 2021; Yilmaz and Sahin, 2021). In general, people who consider the COVID-19 as dangerous and life-threatening are those who are most willing to be vaccinated (Caserotti et al., 2021; Glöckner et al., 2020; Karlsson et al., 2021; Ward et al., 2020). It is possible that these individuals may feel safer through vaccination, not only for themselves but also for their family and friends.

An important parental willingness of vaccination in our study was positive influenza vaccination history in the last season. This finding echoes the results of research which show that COVID-19 vaccine acceptance is higher both among parents who are vaccinated for influenza (Goldman et al., 2020; Kelly et al., 2021) and among parents who vaccinate their children for influenza (Ruggiero et al., 2021). Influenza vaccination shows parents’ positive attitude towards vaccination, which is extremely important as there is a positive relationship between past and future behavior (Ouellette and Wood, 1998). For instance, research has shown the positive relationship between people’s past vaccination and vaccination against the pandemic influenza A/H1N1 vaccine (Bish et al., 2011; Setbon and Raude, 2010; Torun et al., 2010).

The results of our study showed that increased parents’ knowledge regarding COVID-19 was related with decreased parents’ intention to accept a COVID-19 vaccine for their children. High level of positive information about the COVID-19 pandemic is related with parents’ willingness to vaccinate their children (Brandstetter et al., 2021), but higher exposure to negative information increases COVID-19 vaccine hesitancy (Montalti et al., 2021; Zhang et al., 2020). The situation is even more problematic when parents’ information is based on social media (Montalti et al., 2021). Unfortunately, individuals are more likely to absorb negative rather than positive information (Covello, 2003), while major negative vaccine incidents have impaired confidence in vaccines (Liu et al., 2019). In addition, social media in many cases disseminate fake news, unverified rumors and inaccurate data influencing behavioral responses (Seo, 2021). During the COVID-19 pandemic, there has been a global epidemic of misinformation through social media which may have a negative impact on public acceptance of the COVID-19 vaccination programs (Bao et al., 2020; Tangcharoensathien et al., 2020).

Another interesting finding of our study was that increased trust in COVID-19 vaccines and the government regarding the information about the COVID-19 vaccines was associated with parents’ intention to vaccinate their children. Skjefte et al. confirm our finding since they found that trust in public health agencies/health science increases parents’ willingness to vaccinate their children (Skjefte et al., 2021). Citizens’ trust in government is crucial, especially during the COVID-19 pandemic due to the dissemination of fake news in a large extent (Apuke and Omar, 2021; Greene and Murphy, 2021). Misinformation surrounding COVID-19 vaccination threatens public health and could undermine governments’ COVID-19 vaccination programs. Moreover, there is a connection between political populism and parents’ reluctance to vaccinate their children (Kennedy, 2019). Trust in information from government could reduce belief in COVID-19 myths and fake news (Melki et al., 2021). On the other hand, trust in information from social media and interpersonal communication drives individuals to misinformation. Governments must address public fears and misconception using clear messages to create a consensus of the utter importance of COVID-19 vaccination (Skjefte et al., 2021). Also, media literacy training could mitigate the COVID-19 infodemic by increasing critical social media posting practices (Melki et al., 2021).

### Limitations

Our study has a number of limitations. First, we used a convenience sample that is not representative of the general population in Greece. For instance, educational level of our parents is higher than that of the general population. Also, we collected information through social media and parents without social media accounts could not participate in our study. Thus, generalization of our results should be made cautiously. Second, we conducted a cross-sectional study but the situation regarding the COVID-19 vaccination programmes, the dynamic of the COVID-19 pandemic, and the associated policy measures is changing fast. It is therefore necessary to carry out further studies as soon as possible. Indeed, it would be better to carry out prospective studies in order to observe parents’ attitudes over time. Third, since our study was anonymous through social media, we cannot calculate the response rate and we cannot be aware of the profile of parents who refused to participate in the study. Thus, a selection bias is possible since parents who denied to participate in our study might have a different profile as compared to parents that participate. Fourth, the study questionnaire was self-reported and data verification was not feasible. For instance, parents may overestimate their vaccine acceptability due to social desirability. Moreover, we self-constructed some items in our study (e.g. self-perceived severity and knowledge, trust, etc.). Thus, an information bias might exist.

## Conclusions

In conclusion, in September 2021, parents’ intention to vaccinate their children against the COVID-19 was low in our sample of parents in Greece. Also, we found that several factors affect parents’ decision to vaccinate their children. Although preliminary, these findings could contribute to the development of target strategies to implement adherence to COVID-19 vaccination campaigns. Understanding parental vaccine hesitancy is crucial to establish broad community vaccination programmes. There is room for policy makers, scientists, and governments to rebuild confidence in the near future as COVID-19 vaccine roll-out continues.

## Data Availability

Data will be available after a reasonable request

